# Clinical and Radiomics Predictors of Post-stroke Longitudinal Mortality: A Prospective Cohort Study from a Tertiary Care Hospital in India

**DOI:** 10.64898/2026.09.24.26363784

**Authors:** Manju Babu, Sourav Basak, Kathakali Sardar, Mozammil Hassan, Ismail Mohammed Haji, Riyama Shirin, Abhina Gangadhar, Anaekshi Gogoi, Devarati Biswas, Debabrata Chakraborty, Sourav Bhaduri, Arnab Datta

**Affiliations:** Laboratory of Translational Neuroscience, Division of Neuroscience, Yenepoya Research Center, Yenepoya (Deemed to be University), Mangalore 575018, Karnataka, India; Institute for Advancing Intelligence, TCG CREST, Kolkata 700091, West Bengal, India; Department of Radiology, Apollo Multispeciality Hospitals, Kolkata 700054, West Bengal, India; Medical Intensive Care Unit, Yenepoya Medical College & Hospital, Yenepoya (Deemed to be University), Mangalore 575018, Karnataka, India; Department of Pharmacology, Yenepoya Pharmacy College and Research Center, Naringana, Deralakatte, Mangalore 575018, Karnataka, India; Department of Neurology, Apollo Multispeciality Hospitals, Kolkata 700054, West Bengal, India

**Keywords:** Cerebrovascular disorder, Radiomics, Mortality, Prognosis, 1-year Survival, MRI

## Abstract

**Background:** Stroke imposes substantial mortality and socioeconomic burden in India, particularly among younger adults. However, prospective evidence on the prognostic value of baseline clinical and radiomics features for mortality prediction remains limited.

**Objective:** To assess mortality at 28, 90, 180, and 365 days after stroke; characterize survival patterns across stroke subtypes and vascular territories; and identify baseline demographic, clinical, and radiomics predictors of mortality.

**Methods:** This prospective cohort study recruited consecutive patients with stroke admitted to a tertiary care hospital in Mangalore, India. Demographic, clinical, laboratory, and radiomics features were collected at admission, and mortality was assessed at 28, 90, 180, and 365 days. Kaplan–Meier analysis was used to compare survival across stroke subtypes and vascular territories, while Cox proportional hazards regression was used to identify independent baseline predictors of mortality.

**Results:** Among 63 confirmed stroke cases, 38 (60%) had ischemic stroke (IS) and 25 (40%) had haemorrhagic stroke (HS). Within the middle cerebral artery (MCA) territory, HS patients demonstrated better survival than IS patients due to a greater burden of adverse baseline characteristics such as previous stroke, atrial fibrillation, and higher lesion volume in the IS-MCA subgroup. The combined clinical-radiomics model demonstrated superior prognostic performance than individual models, with higher concordance indices and consistently lower Akaike Information Criterion values. In the combined model, stroke severity (aHR = 2.35, 3.0, 2.93, 2.41) and DWI texture heterogeneity of lesion (aHR = 2.35, 2.11, 2.24, 2.10) were the strongest predictors of mortality across all time points. The CT attenuation heterogeneity (aHR = 1.81, 28 days; 1.76, 90 days; 1.80, 180 days) of lesion and age (aHR = 1.48, 365 days) were also significant predictors.

**Conclusion:** The radiomics features provide critical complementary information to clinical parameters for predicting 28-day mortality, unlike during mid- to long-term follow-up where their incremental contribution appears to be limited. Our results may have important implications in resource-constrained settings for supporting personalized post-stroke management and rehabilitation.

## INTRODUCTION

Stroke is a major public health challenge in India and the leading cause of mortality and disability (1). With more than 1.5 million new stroke cases estimated annually, the country carries a substantial and growing stroke burden. Unlike the declining incidence observed in many high-income countries, the burden of stroke has increased considerably in low- and middle-income countries, including India, over the past four decades (1970-2008) (2, 3). Importantly, stroke increasingly affects younger adults in India, resulting in premature mortality, long-term disability, loss of productivity, caregiver dependence, and substantial socioeconomic consequences (4, 5).

With advances in acute stroke care, survival following stroke has increased, especially in younger patients. The surviving patients have a higher risk of having vascular or other complications within the first year, leading to delayed post-stroke mortality (6). Inadequate availability of specialized stroke units, delayed hospital presentation, limited access to reperfusion therapies and rehabilitation centers, and differences in baseline vascular risk profiles contribute to poorer mid- to long-term prognosis (3). Early identification of surviving patients after first-ever stroke, who are at high risk of mortality due to post-stroke complications, is therefore essential for appropriate clinical monitoring, recovery management, and rehabilitation, particularly in resource-constrained settings.

Post-stroke mortality is influenced by several baseline factors, including age, neurological severity, systemic inflammation, comorbidities, stroke subtype, and the extent and distribution of brain injury. Clinical measures such as the National Institutes of Health Stroke Scale (NIHSS) provide important prognostic information; however, they may not fully capture the heterogeneity of cerebral injury. Conventional neuroimaging provides complementary information regarding lesion location and burden, while radiomics enables quantitative characterization of imaging features that may provide additional prognostic information beyond routine visual assessment.

Despite the large and rising stroke burden in India, prospective evidence integrating baseline clinical and quantitative radiomics features to predict mortality over a long-term longitudinal follow-up remains limited. The existing observational studies have largely focused on population-based registries and community-based studies (7), while hospital-based cohort studies are frequently retrospective (8). The prospective studies do not integrate radiomics variables for the determination of long-term mortality (9, 10). Mortality may evolve differently across the early and later post-stroke periods, making assessment across multiple follow-up time points important for understanding the trajectory of survival (9). Further, considering the genetic, geographic, and cultural diversity of India, identifying region-specific mortality predictors may help in fine-tuning health policy and healthcare delivery.

Accordingly, we conducted a prospective longitudinal cohort study of consecutive stroke patients admitted to the Yenepoya Medical College Hospital (YMCH), a tertiary care hospital located in the coastal city of Mangalore, India. We aimed to (1) assess post-stroke mortality at four time points (28, 90, 180, and 365 days); (2) characterize survival patterns across vascular territories and stroke subtypes; and (3) identify baseline clinical and radiomics predictors of mortality. By integrating routinely available clinical characteristics with quantitative radiomics features, this study seeks to improve early identification of patients at higher risk of mortality and provide evidence to support personalised post-stroke management and rehabilitation.

## METHODOLOGY

### Study Population

This study included patients admitted within 14 days of symptom onset with a diagnosis of stroke to the Medical Intensive Care Unit (MICU) of YMCH between August 2024 and June 2025. Participants were eligible for inclusion if they were aged 18 years or older, presented with clinical symptoms compatible with stroke, and had a diagnosis of stroke confirmed using the Trial of Org 10172 in Acute Stroke Treatment (TOAST) criteria (11) through neuroimaging modalities such as non-contrast computed tomography (CT) or magnetic resonance imaging (MRI). Individuals were included irrespective of treatment status, meaning both treated and untreated patients with confirmed stroke were considered eligible for enrolment. Patients with stroke mimics, coagulation disorders, active infection, pregnancy or uncertain pregnancy status in women, traumatic brain injury, or previous neurosurgical intervention were excluded. The detailed exclusion criteria, including specific stroke mimics and medical conditions, are provided in **Supplementary Methods**.

### Data Collection Clinical Data

Demographic and clinical data were collected from the medical records, including age, sex, tobacco use, alcohol use, vascular risk factors (arterial hypertension, diabetes mellitus, dyslipidemia), history of previous stroke, AF, coronary artery disease (CAD), chronic kidney disease (CKD), NIHSS, and Los Angeles Motor Scale (LAMS). Vital signs on arrival, including systolic blood pressure, diastolic blood pressure, and pulse rate, were also recorded. Laboratory findings retrieved from the medical records included blood neutrophil percentage, blood lymphocyte percentage, neutrophil-to-lymphocyte ratio (NLR), sodium, potassium, chloride, C-reactive protein (CRP), Creatine Kinase-MB (CK-MB), and troponin-I levels. In-hospital complications, including pneumonia and post-stroke seizures (PSS), were recorded. Disposition at discharge, including discharge against medical advice (DAMA), was also documented.

### Imaging Protocol and Radiomics Data Acquisition

Patients with stroke symptoms underwent CT examination using a Revolution EVO CT scanner (GE Medical Systems, WI, USA) and MR examination using a clinical 3T MRI scanner (Signa Pioneer, GE Medical Systems, WI, USA) as part of the routine diagnostic procedure. The following sequences were analyzed:

CT was performed with tube voltage = 140 kVp, tube current = 320 mA, exposure time = 1825 ms, exposure = 37 mAs, slice thickness = 1.25 mm (spacing 0.625 mm), reconstruction diameter = 252 mm, gantry tilt = 0°, focal spot size = 1.20 mm, using a SOFT convolution kernel, in a head-first supine position.

Fluid-Attenuated Inversion Recovery (FLAIR) for edema and lesion detection: TR/TE = 9000 / 120.064 ms; TI = 2467.3 ms; flip angle = 160°; slice thickness = 5 mm; pixel spacing = 0.4492 × 0.4492 mm²; matrix size = 512 × 512; FOV = 230 × 230 mm²; orientation = axial; NEX = 1; acquisition time = 3 min 20 s.

Diffusion-Weighted Imaging (DWI) for acute ischemia detection, acquired at two b-values (b = 0 and b = 1000 s/mm²) using spin-echo echo-planar imaging: TR/TE = 5850 / 88.3 ms; flip angle = 90°; slice thickness = 5 mm; pixel spacing = 0.8984 × 0.8984 mm²; matrix size = 256 × 256; phase-encode direction = Anterior to Posterior; NEX = 4; pixel bandwidth = 1304.69 Hz; acquisition time = 3 min 15 s.

### Parameter Maps Extraction

All imaging datasets were anonymized and exported in DICOM format for further processing. CT and FLAIR modalities were co-registered to the DWI sequence using an affine transformation as shown in **Figure 1A-B**. Skull stripping was performed using an automated brain extraction tool, 3D Slicer (v5.8.1). Bias field correction (N4ITK algorithm) was applied to each modality using the SimpleITK Python package (v2.0.2) and Python (v3.7.0) to reduce intensity non-uniformities.

**Figure 1.**
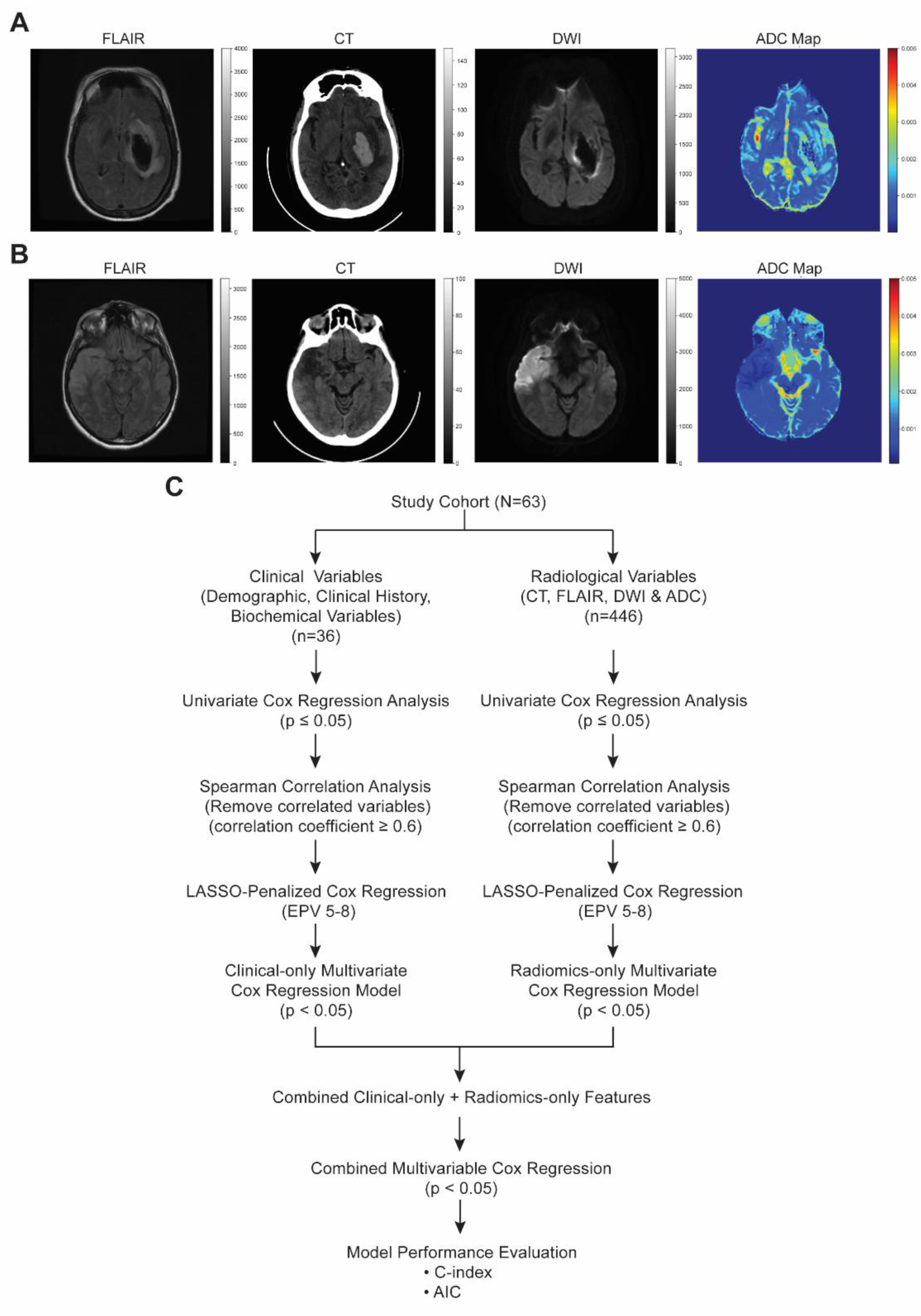
Representative multimodal images of **(A)** a patient with HS and **(B)** and a patient with IS, showing FLAIR and DWI MRI sequences, the ADC map derived from the DWI data, and the corresponding CT scan, with all modalities spatially coregistered to the DWI reference space. **(C)** Workflow for feature selection and development of multivariable Cox regression models, including clinical-only, radiomics-only, and combined clinical-radiomics models.

### Apparent Diffusion Coefficient (ADC) Map Calculation

ADC maps were generated using a mono-exponential decay model from DWI sequences acquired at two b-values (b = 0 and b = 1000 s/mm²). The voxel-wise ADC was computed using the formula:

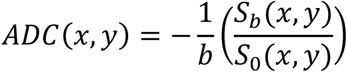

*S*_0_(*x*, *y*) is the signal intensity at b = 0 s/mm²,

*S_b_*(*x*, *y*) is the signal intensity at b = 1000 s/mm²

*b* is the diffusion weighting factor (here, 1000 s/mm²),

(*x*, *y*) denotes the spatial coordinates of each voxel in the image slice.

Stroke lesions were manually delineated by an experienced neuroradiologist on the CT, FLAIR, and DWI images. The segmented lesion Region of Interest (RoI) was mirrored across the longitudinal fissure to obtain a contralateral normal-appearing ROI, used for computing relative signal intensity ratios. From each MRI modality (FLAIR, DWI, ADC map) and CT, quantitative radiomics features were extracted from the segmented lesion areas using the *PyRadiomics* package in Python (v3.1.0).

### Feature Selection

The demographic, clinical, laboratory parameters, radiomics features, and mortality status were included for the analysis. Radiomics features were extracted from the segmented lesion regions of CT, FLAIR, DWI, and ADC images. The extracted radiomics features comprised first-order statistics, shape-based features, Gray Level Co-occurrence Matrix, Gray Level Run Length Matrix, Gray Level Size Zone Matrix, Neighbouring Gray Tone Difference Matrix, and Gray Level Dependence Matrix features. The imaging cohort (N = 63) comprised 21 CT-only, 9 MRI-only, and 33 CT+MRI cases. All available imaging modalities were co-registered to the DWI sequence to ensure spatial correspondence. Missing imaging modality-derived features were addressed using mean imputation (12) based on the available cases.

### Follow-up

All study participants were prospectively followed up at 28, 90, 180, and 365 days by the study team through telephone calls with the caregiver or participant to determine the primary outcome of the study (i.e., mortality status). Participants who could not be reached after repeated telephone attempts were classified as lost to follow-up.

### Statistical Analysis

Data were analysed using IBM SPSS Statistics (v28.0, IBM Corp., Armonk, NY, USA). Continuous variables were assessed for normality using the Shapiro–Wilk test. Normally distributed variables were expressed as mean ± standard deviation (SD), and comparisons between groups were performed using the independent-samples t-test. Non-normally distributed variables were expressed as median and interquartile range (IQR), and group comparisons were conducted using the Mann–Whitney U test. Categorical variables, including sex and vascular risk factors, were presented as frequencies and percentages. Associations between stroke subtype and categorical variables were evaluated using the chi-square test or Fisher’s exact test, as appropriate. RStudio (v2026.05.1, Posit Software, MA, USA) was used for data visualization.

### Survival Analysis

#### Kaplan–Meier Analysis

Survival analysis was performed to evaluate the primary outcome of mortality during the 28, 90, 180, and 365 days follow-up. Patients were stratified according to the anatomical location of the stroke lesion, as determined from neuroimaging findings and manually confirmed by a radiologist blinded to the mortality status or clinical details of the patients. Lesion locations were categorized as middle cerebral artery (MCA), posterior cerebral artery (PCA), anterior cerebral artery (ACA), or multiple locations. Cumulative event curves were constructed using the Kaplan–Meier method. Patients who remained alive at the end of each follow-up period were censored at the corresponding follow-up time, whereas patients lost to follow-up before the end of the study were censored at their last known date of follow-up.

#### Cox Proportional Hazards Regression Model

All variables were first evaluated using univariate Cox regression analyses to identify independent predictors associated with 28, 90, 180 and 365 days survival. Features with a p value of ≤ 0.05 in the univariate analysis were retained for further evaluation **(Figure 1C)**. The multicollinearity among selected features was addressed by pair-wise spearman correlations. For any pair or cluster of highly correlated variables (correlation coefficient ≥ 0.6) (13), the feature with the lower p value in the univariate Cox analysis was retained, and the remaining correlated features were excluded. The remaining clinical variables and radiomics features were considered as candidate predictors for constructing the clinical-only and radiomics-only multivariate Cox regression model. The overfitting risk was minimized by restricting the number of candidate predictors to maintain an events-per-variable (EPV) ratio of approximately 5–8 (14), based on the number of deaths observed at each follow-up time point. Models in which the number of predictors identified through univariate analysis exceeded the recommended EPV ratio, the top six predictors with the non-zero coefficients were retained in the LASSO-penalized Cox regression model (15) for the final multivariate analysis.

#### Model Evaluation

Three multivariate Cox proportional hazards models were developed separately for 28-, 90-, 180- and 365-day mortality: (1) clinical-only model using demographic, clinical, and laboratory variables, (2) radiomics-only model including radiomics features, and (3) combined model including significant features from both clinical and radiomics models. Statistical significance was defined as a p value < 0.05 for every model **Figure 1C**. Adjusted Hazard ratios (aHRs) were reported for all predictors with corresponding 95% confidence intervals (CIs). The aHR > 1 is interpreted as an increased risk of mortality, whereas aHR < 1 as a decreased risk of mortality.

Model selection among the three Cox proportional hazards models (clinical, radiomics, and combined models) were performed using the Akaike Information Criterion (AIC), and the Concordance index (C-index) (16).

The AIC was calculated as follows for each model to evaluate model adequacy while penalizing for complexity,

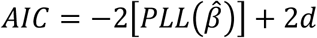

where, d is the effective number of parameters in the model and PLL (*β^*) denotes the maximized partial log-likelihood of *β^*. Lower AIC values reflect a more parsimonious yet better-fitting model.

Finally, prognostic discrimination was quantified using the Concordance, denoted by C-index, which measures the probability that, for a randomly selected pair of individuals, the model correctly orders their predicted risk scores relative to their observed survival times. A higher value of C-index indicates superior discriminative ability. The evaluation criteria AIC and C-index of the three models were compared to establish whether integrating radiomics and clinical predictors improved prognostic performance.

## RESULTS

### Participant Recruitment and Outcome Assessment

Between August 2024 and June 2025, a total of 902 patients admitted to the MICU at YMCH were screened. Of these, 819 patients were excluded because they did not have a neurological diagnosis or severe clinical condition precluding study participation. The remaining 83 patients (9%) presented with stroke-like symptoms and were assessed for eligibility. Among them, 20 were excluded due to suboptimal imaging and medical history suggestive of stroke secondary to a fall, lack of informed consent, or diagnosis of a stroke-mimicking condition. Consequently, 63 patients with confirmed stroke, as diagnosed by the treating clinician and conventional radiological assessment, were included in the outcome analysis. Of these, 60% (38/63) were classified as ischemic stroke (IS) and 40% (25/63) as haemorrhagic stroke (HS) cases. During 1-year follow-up, 41% (26/63), 49% (31/63), 52% (33/63), and 59% (37/63) deaths were recorded by 28, 90, 180, and 365 days from the date of stroke onset. The participant recruitment and follow-up process are illustrated in **Figure 2**.

**Figure 2.**
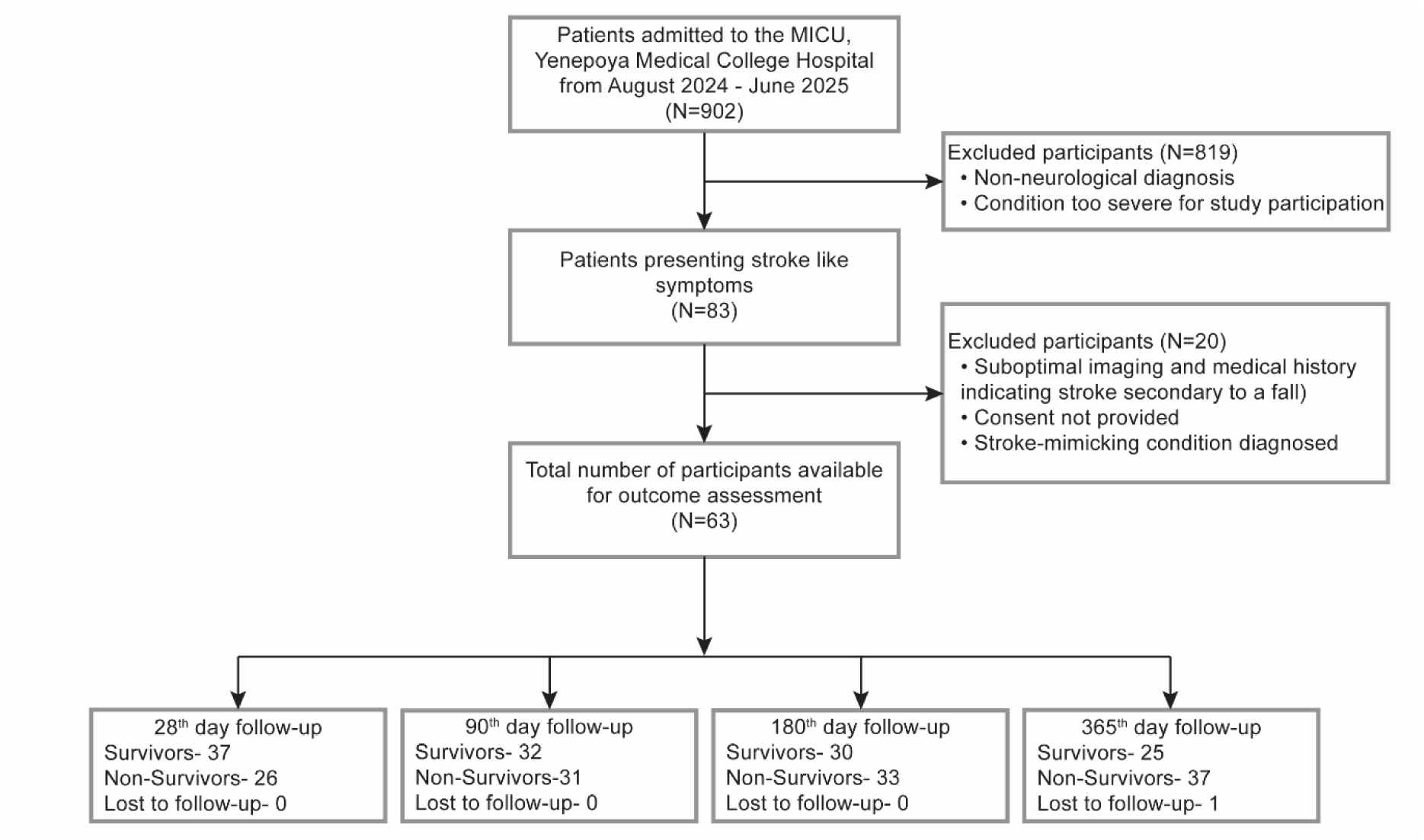
Summary of recruitment and outcome assessment of stroke participants in YMCH at 28, 90, 180, and 365 days. N, number of participants.

### Participant Baseline Characteristics

Baseline demographic and clinical characteristics according to stroke subtype are summarized in **Table 1**. The mean age of patients did not differ significantly between the IS and HS subtypes (59.3 ± 12.7 vs. 55.9 ± 13.9 years, p = 0.330). Sex distribution differed significantly between groups, with males comprising 50.0% of the IS group and 84.0% of the HS group, while females comprised 50.0% and 16.0%, respectively (χ²(1) = 7.52, p = 0.006). Most vascular risk factors, clinical complications, and radiological parameters did not differ significantly between IS and HS patients (p > 0.05). CKD was significantly more common among patients with HS than those with IS (20.0% vs. 2.6%, p = 0.032). Intriguingly, patients with HS had significantly better survival compared to IS at 180 (64.0% vs. 36.8%, χ²(1) = 4.46, p = 0.035) and 365 (62.5% vs. 26.3%, χ²(1) = 8.00, p = 0.005) days. This unusual finding is not explained by higher lesion volume or midline shift in the IS group.

**Table 1.**
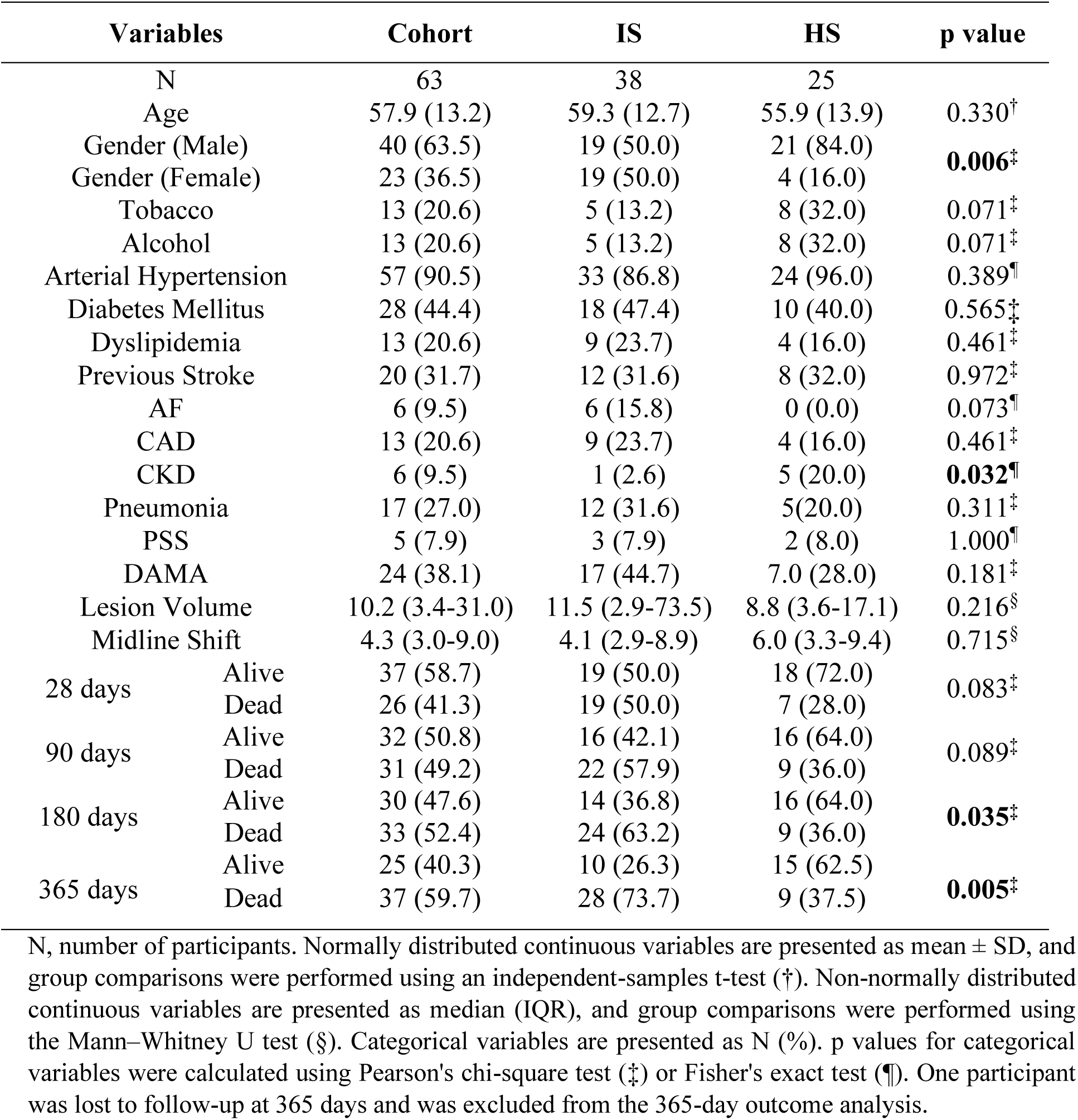
Demographic, clinical, and conventional radiological variables of the patient population.

### Survival Analysis Based on Stroke Type within MCA Territory

The above paradoxical finding in the descriptive statistics **(Table 1)** prompted us to stratify the cohort in terms of affected vascular territories (i.e., MCA, PCA, and ACA). MCA stroke has the maximum prevalence (34/63, 54%), where 17 patients each were in the IS (IS-MCA) and HS (HS-MCA) groups. Kaplan–Meier analysis showed consistently higher cumulative survival in the HS-MCA group than in the IS-MCA group throughout the 1-year follow-up period where at 365 days, the p value was approaching significance (p = 0.054) **(Figure 3)**. Comparison of clinical and conventional radiological parameters between the two MCA stroke groups revealed several significant differences **(Table 2)**. Previous history of stroke was more common in IS-MCA (41.2% vs. 5.9%, p = 0.039). AF was observed among IS-MCA patients but was absent in the HS-MCA group (29.4% vs. 0.0%, p = 0.044). Lesion volume was also significantly greater in IS-MCA than HS-MCA (p = 0.015). Collectively, the higher prevalence of previous stroke and AF, together with greater lesion volume, may contribute to the higher mortality observed in IS-MCA compared with HS-MCA.

**Figure 3.**
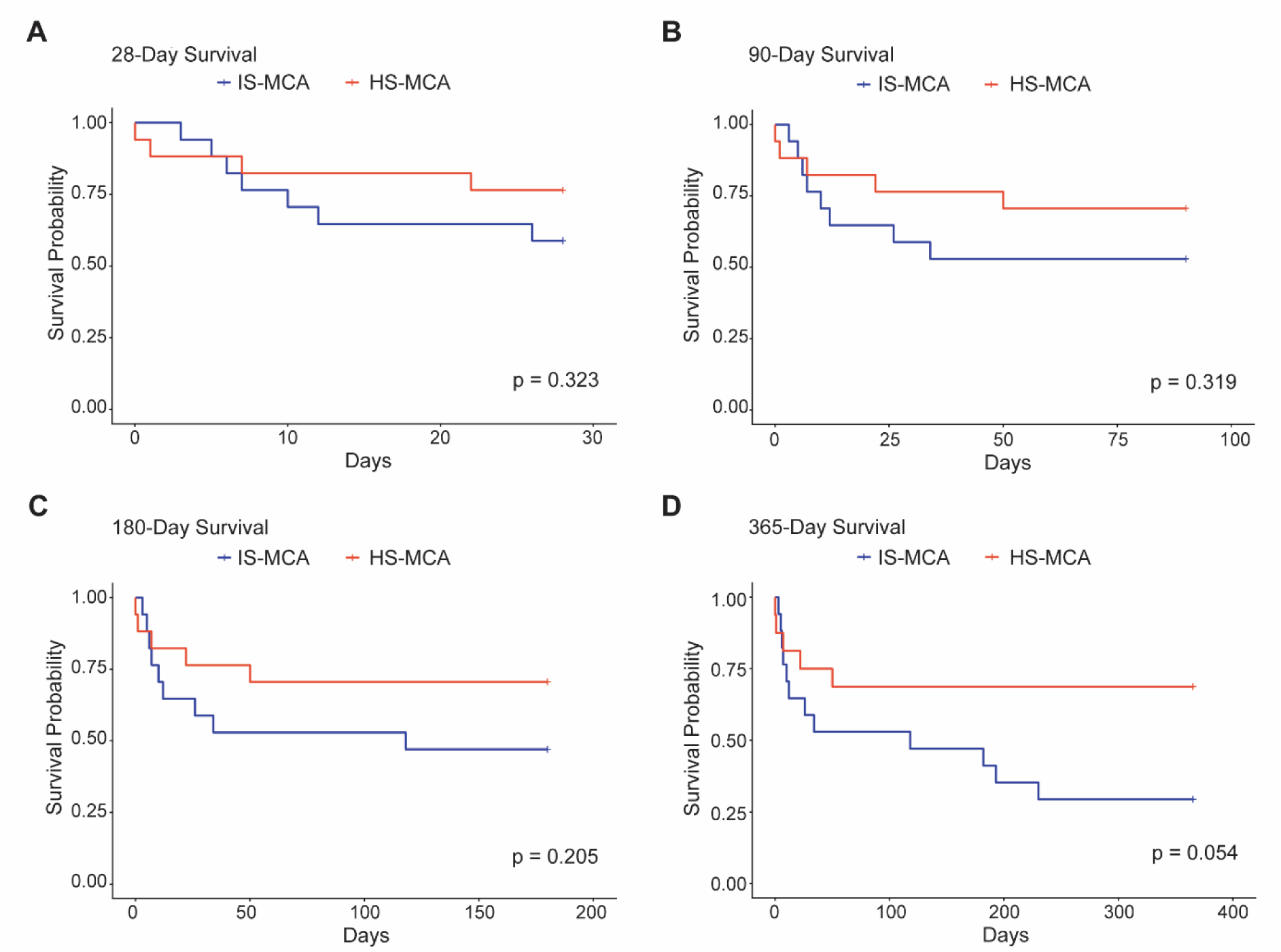
Kaplan–Meier analyses demonstrating cumulative survival probability among patients with IS and HS patients at MCA territory at **(A)** 28 days, **(B)** 90 days, **(C)** 180 days, and **(D)** 365 days.

**Table 2.** Demographic, clinical, and conventional radiological variables of the patients with IS and HS at MCA territory.

| Variables | IS-MCA | HS-MCA | p value |
| --- | --- | --- | --- |
| N | 17 | 17 |  |
| Age (Years) | 59.0 (12.9) | 52.2 (13.4) | 0.140<sup>†</sup> |
| Gender (Male) | 10 (58.8%) | 15 (88.2%) | 0.118<sup>¶</sup> |
| Gender (Female) | 7 (41.2%) | 2 (11.8%) |  |
| Tobacco | 2 (11.8%) | 4 (23.5%) | 0.656<sup>¶</sup> |
| Alcohol | 0 (0.0%) | 5 (29.4%) | **0.044<sup>¶</sup>** |
| Arterial Hypertension | 15 (88.2%) | 16 (94.1%) | 1.000<sup>¶</sup> |
| Diabetes Mellitus | 6 (35.3%) | 7 (41.2%) | 0.724<sup>‡</sup> |
| Dyslipidemia | 4 (23.5%) | 3 (17.6%) | 1.000<sup>¶</sup> |
| Previous Stroke | 7 (41.2%) | 1 (5.9%) | **0.039<sup>¶</sup>** |
| AF | 5 (29.4%) | 0 (0.0%) | **0.044<sup>¶</sup>** |
| CAD | 5 (29.4%) | 3 (17.6%) | 0.688<sup>¶</sup> |
| CKD | 1 (5.9%) | 5 (29.4%) | 0.175<sup>¶</sup> |
| Pneumonia | 5 (29.4%) | 4 (23.5%) | 1.000<sup>¶</sup> |
| PSS | 1 (5.9%) | 2 (11.8%) | 1.000<sup>¶</sup> |
| DAMA | 7 (41.2%) | 5 (29.4%) | 0.473<sup>‡</sup> |
| Lesion Volume | 40.2 (7.0-127.9) | 6.7 (3.6- 17.1) | **0.015<sup>§</sup>** |
| Midline Shift | 4.3 (2.8-11.6) | 6.5 (2.8-11.5) | 0.783<sup>§</sup> |

N, number of participants. Normally distributed continuous variables are presented as mean ± SD, and group comparisons were performed using an independent-samples t-test (<sup>†</sup>). Non-normally distributed continuous variables are presented as median (IQR), and group comparisons were performed using the Mann–Whitney U test (<sup>§</sup>). Categorical variables are presented as N (%). P values for categorical variables were calculated using Pearson's chi-square test (<sup>‡</sup>) or Fisher's exact test (<sup>¶</sup>).

### Multivariate Cox Regression using Clinical Features

Clinical variables identified as candidate predictors through univariate analysis were used as selected features for constructing four independent multivariate Cox proportional hazards regression models to identify independent predictors of mortality at 28, 90, 180, and 365 days. The results of the univariate analyses are provided in the **Supplementary Results (Supplementary Table 2)**. The NIHSS was consistently and significantly associated with mortality across all follow-up periods, with aHRs of 1.07, 2.57, 2.30, and 2.09 at 28, 90, 180, and 365 days, respectively. Similarly, NLR demonstrated significant positive associations with mortality, with aHRs of 1.07, 2.17, 1.93, and 1.92 across the respective follow-up periods. Age also showed a strong positive association with mortality during 90 – 365 days, with highest magnitude of association observed at 6 months (aHRs: 1.05, 28 days; 2.20, 90 days; 2.21, 180 days; and 1.97, 365 days). Overall, the clinical predictors showed stronger associations with mid-to-long-term mortality (i.e., 90, 180, and 365 days) compared with early mortality (i.e., 28 days) **(Table 3**, **Figure 4A)**.

**Figure 4.**
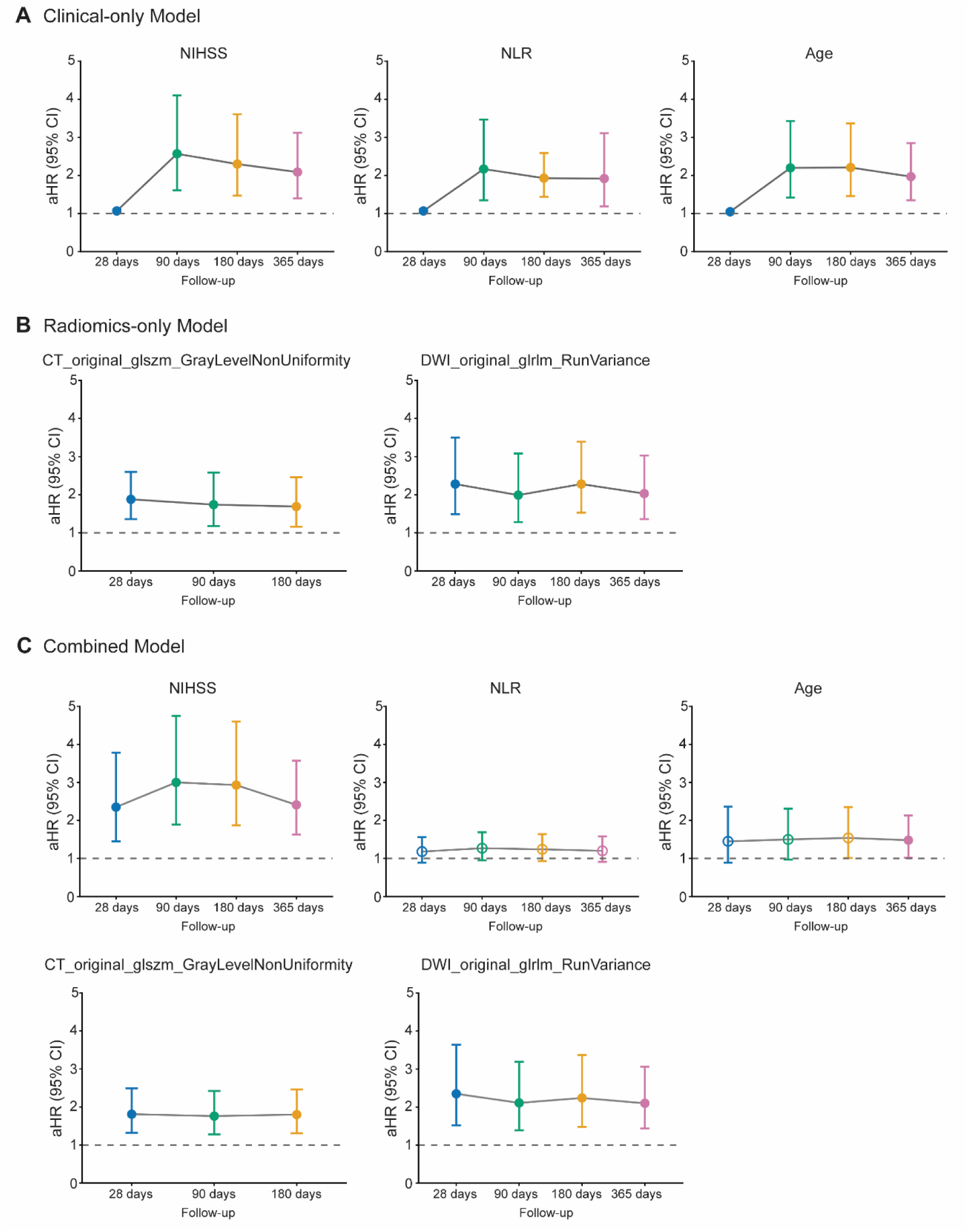
Multivariate Cox regression analysis of predictors of mortality across follow-up. aHRs with 95% CIs are shown for significant predictors at 28, 90, 180, and 365 days. **(A)** Clinical-only predictors; **(B)** Radiomics-only predictors; and **(C)** Combined clinical and radiomics predictors identified by multivariate Cox regression analysis. The dashed horizontal line represents aHR = 1, indicating no association with mortality. All predictors shown were statistically significant (p < 0.05).

**Table 3.** Features from multivariate clinical model.

| Features | aHR (95% CI) | p value |
| --- | --- | --- |
| **28 days** |  |  |
| NIHSS | 1.07 (1.03-1.11) | <0.005* |
| NLR | 1.07 (1.02-1.12) | 0.005* |
| Age (years) | 1.05 (1.01-1.08) | 0.009* |
| CRP (mg/L) | 1.00 (0.99-1.01) | 0.458 |
| **90 days** |  |  |
| NIHSS | 2.57 (1.61-4.10) | <0.005* |
| NLR | 2.17 (1.35-3.47) | <0.005* |
| Age (years) | 2.20 (1.42-3.43) | <0.005* |
| Stroke Onset-to-Admission Time (days) | 1.46 (0.90-2.35) | 0.124 |
| Gender | 0.57 (0.25-1.30) | 0.180 |
| CRP (mg/L) | 0.78 (0.46-1.32) | 0.349 |
| **180 days** |  |  |
| NIHSS | 2.30 (1.47-3.61) | <0.005* |
| Age (years) | 2.21 (1.46-3.37) | <0.005* |
| NLR | 1.93 (1.44-2.59) | <0.005* |
| Gender | 0.58 (0.26-1.32) | 0.196 |
| Diagnosis | 1.79 (0.69-4.68) | 0.235 |
| Stroke Onset-to-Admission Time (days) | 1.27 (0.82-1.98) | 0.289 |
| **365 days** |  |  |
| NIHSS | 2.09 (1.40-3.12) | <0.005* |
| Age (years) | 1.97 (1.35-2.85) | <0.005* |
| NLR | 1.92 (1.19-3.11) | 0.007* |
| Diagnosis | 2.45 (0.96-6.23) | 0.060 |
| Gender | 0.55 (0.27-1.12) | 0.100 |
| CRP (mg/L) | 0.97 (0.61-1.54) | 0.881 |

\*p < 0.05, statistically significant

### Multivariate Cox Regression using Radiomics Features

Radiomics variables identified as candidate predictors through univariate analysis were subsequently used as selected features for constructing four independent multivariate Cox proportional hazards regression models to identify independent predictors of mortality at 28, 90, 180, and 365 days. The results of the univariate analyses are provided in the **Supplementary Results (Supplementary Table 3)**. CT_original_glszm_GrayLevelNonUniformity was significantly associated with mortality at 28, 90, and 180 days, with aHRs of 1.88, 1.74, and 1.69, respectively. DWI_original_glrlm_RunVariance demonstrated a significant association across all follow-up periods, with aHRs of 2.28, 1.99, 2.28, and 2.03 at 28, 90, 180, and 365 days, respectively. These findings indicate that higher values of both radiomics features were independently associated with increased mortality hazard. Strikingly, unlike clinical predictors, the radiomics features demonstrated their strongest associations with early mortality at 28 days, with broadly consistent effects across subsequent follow-up periods **(Table 4**, **Figure 4B)**.

**Table 4.** Features from multivariate radiomics model.

| Features | aHR (95% CI) | p value |
| --- | --- | --- |
| **28 days** |  |  |
| CT_original_glszm_GrayLevelNonUniformity | 1.88 (1.36-2.60) | <0.005* |
| DWI_original_glrlm_RunVariance | 2.28 (1.49-3.50) | <0.005* |
| DWI_original_firstorder_Skewness | 1.21 (0.87-1.68) | 0.256 |
| CT_diagnostics_Image.original_Maximum | 1.25 (0.85-1.85) | 0.265 |
| **90 days** |  |  |
| DWI_original_glrlm_RunVariance | 1.99 (1.28-3.08) | <0.005* |
| CT_original_glszm_GrayLevelNonUniformity | 1.74 (1.18-2.58) | 0.006* |
| DWI_original_firstorder_Skewness | 1.19 (0.85-1.68) | 0.310 |
| FLAIR_original_firstorder_TotalEnergy | 1.24 (0.78-1.99) | 0.366 |
| CT_diagnostics_Image.original_Maximum | 1.15 (0.75-1.77) | 0.512 |
| CT_original_shape_MinorAxisLength | 1.04 (0.64-1.69) | 0.880 |
| **180 days** |  |  |
| DWI_original_glrlm_RunVariance | 2.28 (1.53-3.39) | <0.005* |
| CT_original_glszm_GrayLevelNonUniformity | 1.69 (1.16-2.46) | 0.006* |
| CT_original_shape_MinorAxisLength | 1.21 (0.78-1.87) | 0.401 |
| CT_diagnostics_Image.original_Maximum | 1.14 (0.79-1.65) | 0.481 |
| **365 days** |  |  |
| DWI_original_glrlm_RunVariance | 2.03 (1.36-3.03) | <0.005* |
| CT_original_shape_MinorAxisLength | 1.51 (0.99-2.29) | 0.053 |
| DWI_original_firstorder_Skewness | 1.12 (0.84-1.51) | 0.441 |
| CT_diagnostics_Image.original_Maximum | 1.08 (0.76-1.54) | 0.670 |
| FLAIR_original_firstorder_Kurtosis | 0.98 (0.66-1.47) | 0.928 |

\*p < 0.05, statistically significant

### Multivariate Cox Regression using Significant Clinical and Radiomics Features

In the combined multivariate model constructed using three significant clinical and two radiomics predictors, NIHSS demonstrated a consistent and statistically significant association with mortality, with aHRs of 2.35, 3.00, 2.93, and 2.41 at 28, 90, 180, and 365 days, respectively. The strongest association in mortality hazard was observed at 3-month **(Table 5**, **Figure 4C)**. Age also showed a positive consistent association, with aHRs of 1.45, 1.50, 1.54, and 1.48 across the four follow-up periods. However, statistical significance was observed only at 365 days. Among the radiomics variables, CT_original_glszm_GrayLevelNonUniformity was significantly associated with mortality at 28, 90, and 180 days, with aHRs of 1.81, 1.76, and 1.80, respectively. In contrast, DWI_original_glrlm_RunVariance showed a significant association across all follow-up periods, with aHRs of 2.35, 2.11, 2.24, and 2.10, respectively. Overall, NIHSS and DWI_original_glrlm_RunVariance emerged as the most consistent independent predictors of mortality followed by CT_original_glszm_GrayLevelNonUniformity and age.

**Table 5.** Multivariate Cox regression analyses using clinical and radiomics features.

| Features | aHR (95% CI) | p value |
| --- | --- | --- |
| **28 days** |  |  |
| Age | 1.45 (0.89-2.36) | 0.130 |
| NIHSS | 2.35 (1.45-3.78) | <0.005* |
| NLR | 1.18 (0.89-1.56) | 0.250 |
| CT_original_glszm_GrayLevelNonUniformity | 1.81 (1.32-2.49) | <0.005* |
| DWI_original_glrlm_RunVariance | 2.35 (1.52-3.64) | <0.005* |
| **90 days** |  |  |
| Age | 1.50 (0.97-2.31) | 0.060 |
| NIHSS | 3.00 (1.89-4.75) | <0.005* |
| NLR | 1.27 (0.95-1.69) | 0.100 |
| CT_original_glszm_GrayLevelNonUniformity | 1.76 (1.28-2.42) | <0.005* |
| DWI_original_glrlm_RunVariance | 2.11 (1.39-3.19) | <0.005* |
| **180 days** |  |  |
| Age | 1.54 (1.01-2.35) | 0.050 |
| NIHSS | 2.93 (1.87-4.60) | <0.005* |
| NLR | 1.24 (0.93-1.64) | 0.140 |
| CT_original_glszm_GrayLevelNonUniformity | 1.80 (1.31-2.46) | <0.005* |
| DWI_original_glrlm_RunVariance | 2.24 (1.48-3.37) | <0.005* |
| **365 days** |  |  |
| Age | 1.48 (1.02-2.13) | 0.040* |
| NIHSS | 2.41 (1.63-3.57) | <0.005* |
| NLR | 1.20 (0.91-1.58) | 0.210 |
| DWI_original_glrlm_RunVariance | 2.10 (1.44-3.06) | <0.005* |

\*p < 0.05, statistically significant

### Model Performance

The combined clinical-radiomics model consistently demonstrated the highest discriminative ability and the best relative model fit across all follow-up periods, as indicated by higher C-index values and lower AIC values compared with the clinical-only and radiomics-only models. Notably, radiomics-only model showed better discrimination (i.e., higher C-index) than clinical-only model at 28 days. However, at all subsequent time-points, clinical-only model performed better compared to radiomics-only model for C-index and AIC values. Further, the incremental benefit of radiomics features was most pronounced for 28-day outcome, with an improvement in C-index from 0.76 for the clinical model to 0.86 for the combined model (ΔC-index = 0.10) and a ΔAIC of 18.2 **(Table 6)**. This indicates that for predicting early mortality (i.e., 28 days), radiomics parameters are particularly informative. A gradual decline in both the ΔC-index and ΔAIC (28 – 365 days) suggests that although radiomics features add meaningful prognostic information beyond clinical variables during 28-180 days, the magnitude of their incremental contribution decreases at longer follow-up (i.e., 365 days) **(Table 6)**.

**Table 6.**
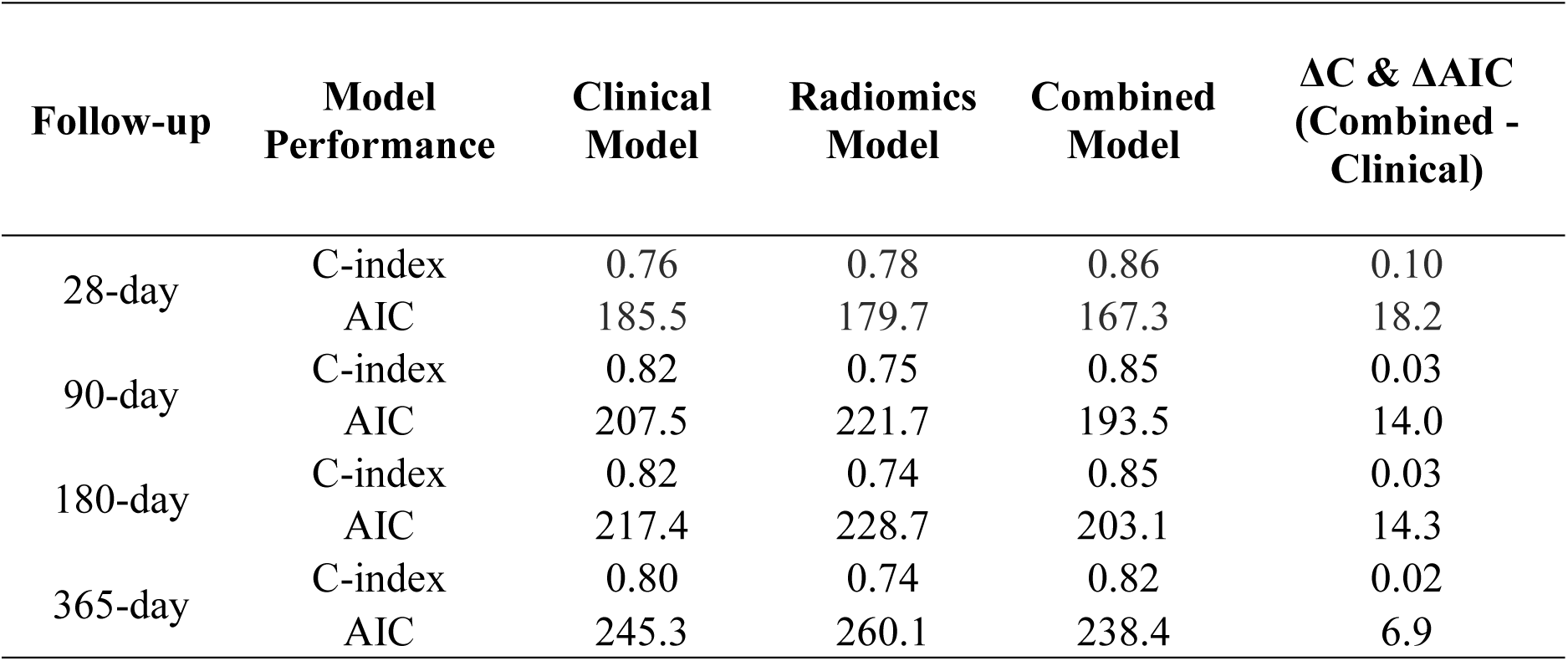
Model performance using clinical, radiomics, and combined features.

## DISCUSSION

India bears a substantial and growing burden of stroke, with an increasing proportion occurring among younger adults creating a large population of surviving stroke patients. In our study, 8% (5/63) and 29% (18/63) of the affected individuals were younger than 35 and 50 years. The prospective design and serial follow-up of radiologically confirmed stroke cases enabled a comprehensive early (28 days), intermediate (3-6 months) and long-term (1 year) assessment of the post-stroke mortality and its baseline predictors. A substantial proportion of deaths (70%, 26/37) occurred within the first 4-weeks, indicating that the early post-stroke period represents the most vulnerable phase for the recovering patients. A detailed analysis of case records in this cohort showed that an unusually large percentage of patients (38%, 24/63) were discharged against medical advice in a critical condition, majority of whom died before the first follow-up (i.e., 28 days). Further, a majority (>50%) of the survived patients did not come back for the mandatory follow-up (e.g., 4-week, 3, or 6 month) visits. This could be due to a combination of multiple factors such as shifting to a different healthcare facility, worsening health with progressively increasing morbidity, financial or logistic constraints, or a general lack of awareness about the grave prognosis, or complete recovery. This further justifies the necessity of identifying predictors of long-term survival from the baseline clinical and radiomics variables that can be collected during the hospital stay. This way, it will be possible to counsel the patient or caregiver before discharge about the possible risk of adverse outcome and advice on optimum rehab paradigm and secondary prevention strategies.

This study built a predictive model for mortality evaluation by combining baseline demographic, clinical, laboratory, and multimodal radiomics features on Indian stroke patients for the first time to determine a temporal profile for significant predictors. Among the clinical variables, NIHSS, age, and NLR emerged as consistent independent predictors of mortality across all follow-up periods, highlighting the sustained prognostic importance of functional neurological impairment, age-related vulnerability, and systemic inflammation after stroke **(Figure 4A**, **Table 3)**. NIHSS showed the strongest and most consistent association, supporting previous evidence that baseline stroke severity is a major determinant of both short- and long-term mortality (9, 17). The persistent association of age with mortality may reflect reduced physiological reserve, greater comorbidity burden, and limited recovery capacity in older patients (18, 19). NLR remained independently associated with mortality even after adjustment for age and NIHSS, suggesting that systemic inflammatory status provides prognostic information beyond conventional clinical measures and may serve as an accessible adjunct for long-term risk stratification (20, 21).

The texture-based radiomics features derived from conventional imaging provide independent prognostic information for mortality following IS. Among all extracted radiomics descriptors, features reflecting diffusion and CT texture heterogeneity demonstrated the most consistent associations with mortality. In particular, DWI_original_glrlm_RunVariance, which represents spatial heterogeneity of the diffusion signal within the lesion remained independently associated with mortality across all follow-up periods, while CT_original_glszm_GrayLevelNonUniformity, reflecting heterogeneity of CT attenuation within the lesion remained independently associated with short- (28 days) and intermediate-term (90 and 180 days) mortality **(Figure 4B**, **Table 4)**. This indicates that quantification of intralesional imaging heterogeneity captures clinically relevant information beyond lesion morphology or first-order intensity features. The predominance of texture-based features in our study is consistent with the growing number of radiomics studies in stroke, in which texture-based features have been shown to be of greater prognostic value than shape-or intensity-based features (22–24). Many studies have shown that texture-based features derived from CT and MRI scans can predict functional outcome, infarct progression, and response to therapy (22–24). However, the choice of radiomics features depends heavily on the imaging techniques used and the study endpoint (22, 23).

After integrating radiomics-based features into the clinical combination model, DWI_original_glrlm_RunVariance was significantly associated with mortality, even after adjusting for other known clinical variables such as NIHSS, age, and NLR **(Figure 4C)**. While NIHSS was found to be the most powerful clinical predictor across all follow-up periods, the consistently significant value of DWI texture heterogeneity indicates that imaging conveys information beyond neurological deficits or systemic inflammation alone. Importantly, NLR was no longer an independent predictor of mortality after the inclusion of radiomics features in the model. Thus, radiomics markers appear to have complementary predictive value when compared with traditional laboratory-based indicators such as NLR. The results are consistent with earlier research showing that integrating clinical and radiomics features can improve prognostic accuracy compared with clinical models alone (22, 23). However, our findings also have important implications for resource-constrained settings in the low-to-middle income countries, where access to advanced imaging and computational infrastructure may be limited or when patient does not have baseline neuroimaging details.

The good prognostic performance achieved using clinical features alone especially during 90 – 365 days suggests that stroke outcomes can be reasonably predicted using routinely available demographic and clinical parameters without requiring additional radiomics analysis **(Table 6)**. Thus, while radiomics may further refine mortality prediction particularly during early phase (28 days, ΔC = 0.10, ΔAIC = 18.2, **Table 6**). Overall, it should be considered a complementary rather than essential component of practical and cost-effective stroke prognostication.

Our study has several limitations. First, we could not confirm the completeness of the clinical history that was manually collected from the accompanying caregiver without supporting clinical reports in many cases. It is possible that the prevalence of comorbid conditions (e.g., dyslipidemia, hypertention, ischemic heart disease, previous stroke) may have been underestimated due to a reporting bias stemming from a general lack of awareness among the study population. Second, nearly half of the patients (CT-only, 21; MRI-only, 9; 30/63) in our cohort did not have complete multimodal imaging data available. The is because the decision to perform CT or MRI depends on the clinical necessity and there are situations where only CT (e.g., HS) or only MRI (e.g., lacunar stroke) may be sufficient for patient management. We have addressed this by using mean imputation technique (12). Third, we have not collected any functional data beyond the mortality status during the 1-year telephonic follow-up. This was done purposefully as we cannot verify the quality and consistency of functional information coming from the caregivers who might change during the follow-up period. Thus, alive does not mean the patients have recovered or functionally normal. It is likely that several patients who are alive after one year are severely disabled (i.e., modified Rankin scale = 4-5), bedridden, and under palliative care. In some cases (n = 4), we could not obtain the exact date of death via telephone call and used 28-day during survival analysis. This can overestimate the number of days the patient survived. To rule out any systematic bias, we re-analysed the survival curve using the anticipated minimum number of survival days and found no change in the final trends. Fourth, our study has a small sample size (n = 63) despite screening 902 patients. However, it should be noted that for the feasibility of Cox regression model, event (i.e., death) count is more important than the total number of recruited subjects. To the best of our knowledge, case fatality of our cohort is probably the highest (37/63, 59%) among all reported cohort studies on stroke. Using an EPV count of 5-8, we have carefully selected five predictors to build the combined multivariate model to achieve a balance between simplicity and predictive accuracy. Fifth, different types of biases can explain this disproportionately high mortality in our cohort. 1) Severity bias: the patients are primarily recruited from the emergency department of a tertiary care hospital. In general, hospital-based studies is known to have higher event (i.e., death) rates compared to population-based cohorts as patients who cannot be treated at home, reach hospital for treatment. Further, emergency department deals with critical cases with serious complications compared to outpatient department that generally deals with mild cases (25). Hence, critically-ill stroke patients are overrepresented in our cohort (NIHSS >25, 23/63 = 37%). 2) Referral bias: the study site (i.e., YMCH) is a tertiary care center, where complicated or untreatable cases are referred from the peripheral centers. The median time of presentation after stroke onset is 1 days indicating a referral delay. Hence, mild cases (NIHSS <5, 8/63 =13%) are underrepresented in our cohort. 3) Socioeconomic access bias: treatments are subsidized at YMCH through government insurance schemes. Hence, poor and socially underprivileged population are likely overrepresented in this cohort. Unfortunately, we have not collected data on religion, education, and occupation.

## CONCLUSION

Mortality was highest during the acute phase of stroke, particularly within the first 28 days. In the MCA territory, a higher lesion volume alongside baseline risk factors such as AF, previous stroke can make IS more severe than HS. Combined multivariable analysis identified one clinical (NIHSS), and two radiomics features (i.e., CT_original_glszm_GrayLevelNonUniformity, and DWI_original_glrlm_RunVariance) as the most consistent independent predictors of mortality, highlighting the complementary prognostic value of stroke severity and imaging-derived radiomics features. Although integrating radiomics features with clinical variables improve risk prediction during early phase, the good performance of clinical features alone during mid-to-long term phase demonstrates that routine clinical parameters such as NIHSS, NLR, and age can provide effective prognostic information in resource-constrained settings. Radiomics therefore represents a valuable complementary tool rather than an essential requirement for practical stroke prognostication.

## Supplementary Information

The supplementary material is available in the supplementary file accompanying this article.

## Data Availability

All data produced in the present study are available upon reasonable request to the authors.

## Acknowledgements

The authors thank Mashood C, Shaymah Abdurahiman, and Deepthi Ann Thomas for assistance during patient recruitment and data collection at the YMCH.

## Author Contributions

Conceptualization: A.D.; Study Design: A.D., I.M.H.; Funding Acquisition: A.D., S.B.; Data collection: M.B., S.Ba, K.S., M.H., R.S., A.Ga., A.Go., H.M.I., S.B., A.D; Data Analysis: M.B., S.Ba., K.S., M.H., S.B., A.D., D.C., D.B.; Writing - Original Draft: M.B., S.Ba, K.S., S.B., A.D.; Writing – Review & Editing: A.D. All authors approved the final version of the manuscript.

## Funding

Arnab Datta acknowledges funding support from Vision Group on Science and Technology Infrastructure Grant (K-FIST(L2)/2024, GRD No. 1128), Department of Science & Technology, Govt. of Karnataka and an institutional seed grant (YU/Seed Grant/169-2024) at Yenepoya (Deemed to be University). Manju Babu was funded by a senior research fellowship under the guidance of Arnab Datta from ICMR (BMI/11(67)/2022). Abhina Gangadhar is funded by a junior research fellowship from CSIR under the supervision of Arnab Datta (File No. 08/0652(19079)/2024-EMR-I). Riyama Shirin and Anaekshi Gogoi are funded by a junior research fellowship from Yenepoya (deemed to be University). Sourav Bhaduri acknowledges the Ramalingaswami Re-entry Fellowship (BT/RLF/Re-entry/37/2022) from the Department of Biotechnology (DBT), Government of India, for supporting the computational resources used in the radiomics analysis, the research fellowship of Sourav Basak, and the internship stipend of Kathakali Sardar.

## Data availability

The data that support the findings of this study are available from the corresponding authors upon reasonable request.

## DECLARATIONS

### Conflict of Interest

On behalf of all authors, the corresponding author states that there is no conflict of interest.

### Ethics Statement

The study was conducted in accordance with the Declaration of Helsinki (2013) and approved by the Yenepoya Ethics Committee-1 of Yenepoya (Deemed to be University) (Approval No. YEC-1/2024/200). A Data Usage Agreement was signed between Yenepoya (Deemed to be University) and TCG Centres for Research and Education in Science and Technology (TCG CREST) for sharing the clinical and imaging data.

### Informed Consent

Written informed consent was obtained from all participants or their legally authorized representatives prior to inclusion in the study.

### Abbreviations

IS: Ischemic stroke
HS: Hemorrhagic stroke
NIHSS: National Institutes of Health Stroke Scale
YMCH: Yenepoya Medical College Hospital
MICU: Medical intensive care unit
CT: Computed tomography
MRI: Magnetic resonance imaging
AF: Atrial fibrillation
CAD: Coronary artery disease
CKD: Chronic kidney disease
LAMS: Los Angeles Motor Scale
NLR: Neutrophil-to-lymphocyte ratio
CRP: C-reactive protein
CK-MB: Creatine kinase-MB
PSS: Post-stroke Seizures
DAMA: Discharge Against Medical Advice
FLAIR: Fluid-attenuated inversion recovery
DWI: Diffusion-weighted imaging
ADC: Apparent diffusion coefficient
EPV: Events-per-variable
aHRs: Adjusted hazard ratios
CIs: Confidence intervals
AIC: Akaike Information Criterion
C-index: Concordance index
MCA: Middle cerebral artery
PCA: Posterior cerebral artery
ACA: Anterior cerebral artery
RoI: Region of Interest
SD: Standard deviation
IQR: Interquartile range

## Notes

### Competing Interest Statement

The authors have declared no competing interest.

