## Supplementary File for "Clinical and Radiomics Predictors of Post-stroke Longitudinal Mortality: A Prospective Cohort Study from a Tertiary Care Hospital in India"



<u>**Index**</u>


**Supplementary Methods**

Exclusion Criteria

**Supplementary Results**

Baseline Characteristics of Survivors and Non-Survivors Over Time

Univariate Cox Regression using Clinical Features

Univariate Cox Regression using Radiomics Features

**Supplemental Tables**

**Supplementary Table 1.** Demographic and clinical characteristics of survivors and non-survivors at 28, 90, 180, and 365 days

**Supplementary Table 2.** Clinical features obtained from univariate Cox regression analyses

**Supplemental Table 3.** Radiomics features obtained from univariate Cox regression analyses

### Supplementary Methods

#### Exclusion Criteria

Stroke mimetics considered for exclusion included seizures, secondary metastatic lesions, sepsis, migraine, severe headache, postictal hemiparesis, otogenic vertigo, or transient focal neurological deficits resulting from systemic metabolic disturbances. Individuals with known coagulation disorders, such as hemophilia, clotting factor deficiencies, von Willebrand disease, or deep vein thrombosis, were also excluded. Additional exclusion criteria included the presence of any active bacterial, viral, or fungal infection at the time of admission, pregnancy or uncertain pregnancy status in females younger than 50 years, a history of traumatic brain injury, or prior neurosurgical intervention.

### Supplementary Results

#### Baseline Characteristics of Survivors and Non-Survivors Over Time

Across all follow-up periods, non-survivors were consistently older than survivors, with significant differences observed at 28 days (63.2 vs. 54.2 years, $p = 0.007$), 90 days (62.9 vs. 53.2 years, $p = 0.003$), 180 days (64.0 vs. 53.0 years, $p = 0.002$), and 365 days (61.5 vs. 52.1 years, $p = 0.005$). These findings indicate that increasing age was strongly associated with mortality throughout the follow-up period. Sex distribution also demonstrated an increasingly significant association with survival over time. Although the difference was not statistically significant at 28 days ($p = 0.062$) and borderline at 90 days ($p = 0.054$), male patients constituted a significantly higher proportion of survivors at 180 days (80.0% vs. 48.5%, $p = 0.009$) and 365 days (84.0% vs. 48.6%, $p = 0.005$). Most vascular risk factors, including tobacco use, alcohol consumption, arterial hypertension, diabetes mellitus, dyslipidemia, AF, and CKD, did not differ significantly between survivors and non-survivors at any time point ($p > 0.05$). However, previous stroke was significantly more common among non-survivors at 90 days (45.2% vs. 18.8%, $p = 0.024$) and remained higher at 180 days (42.4% vs. 20.0%, $p = 0.056$) and 365 days (40.5% vs. 20.0%, $p = 0.090$), suggesting a potential association with poorer long-term outcomes. CAD emerged as a significant predictor of long-term mortality, showing no significant association at earlier time points but becoming significantly more prevalent among non-survivors at 365 days (29.7% vs. 8.0%, $p = 0.039$). Overall, increasing age was the most consistent factor associated with mortality across all follow-up periods, while previous stroke and CAD appeared to contribute predominantly to longer-term mortality risk (**Supplementary Table 1**).

**Univariate Cox Regression using Clinical Features**

A total of 36 clinical features involving demographic, clinical history, and biochemical variables were initially evaluated using univariate Cox proportional hazards regression to shortlist the predictors of mortality at 28, 90, 180, and 365 days following stroke **(Supplementary Table 2)**. At 28 days, higher NIHSS scores (HR: 1.06, $p < 0.005$), NLR (HR: 1.04, $p < 0.005$), age (HR: 1.05, $p = 0.011$), and CRP levels (HR: 1.01, $p = 0.037$) were significantly associated with an increased risk of mortality. At 90 days, NIHSS remained the strongest predictor of mortality (HR: 2.34, $p < 0.005$), followed by NLR, CRP, age, stroke onset-to-admission time, and LAMS score, all of which were associated with an increased mortality risk. In contrast, higher hemoglobin (HR: 0.64, $p = 0.024$), albumin (HR: 0.70, $p = 0.037$), and male patients (HR: 0.48, $p = 0.042$) were associated with a lower risk of mortality. At 180 days, higher NIHSS scores (HR: 2.28, $p < 0.005$), NLR (HR: 1.52, $p < 0.005$), age (HR: 1.87, $p < 0.005$), CRP (HR: 1.56, $p < 0.005$), stroke onset-to-admission time (HR: 1.61, $p < 0.005$), LAMS score (HR: 1.54, $p = 0.035$), and diagnosis (IS/ HS) (HR: 2.17, $p = 0.048$) were significantly associated with an increased risk of mortality. Conversely, higher hemoglobin (HR: 0.63, $p = 0.017$), higher albumin (HR: 0.70, $p = 0.031$), and male patients (HR: 0.41, $p = 0.012$) were associated with a reduced risk of mortality. At 365 days, higher NIHSS scores (HR: 2.15, $p < 0.005$), CRP (HR: 1.58, $p < 0.005$), age (HR: 1.73, $p < 0.005$), NLR (HR: 1.46, $p = 0.005$), stroke onset-to-admission time (HR: 1.57, $p = 0.008$), and diagnosis (HR: 2.54, $p = 0.016$) remained significantly associated with an increased risk of mortality. In contrast, male patients (HR: 0.40, $p = 0.005$), higher albumin (HR: 0.64, $p = 0.006$), and higher hemoglobin levels (HR: 0.61, $p = 0.008$) remained significantly associated with reduced mortality risk. Across all follow-up periods, NIHSS, NLR, age, and CRP consistently emerged as significant predictors of mortality, whereas higher hemoglobin and albumin concentrations, along with male sex, demonstrated protective associations during longer-term follow-up.

**Univariate Cox Regression using Radiomics Features**

A total of 446 radiomics features were extracted from the segmented lesion regions of each imaging modality (CT, FLAIR, DWI, and ADC) and evaluated for their association with mortality. The extracted feature set comprised 19 first-order statistics, 26 shape-based features, 24 GLCM features, 16 GLRLM features, 16 GLSZM features, 5 NGTDM features, and 14

GLDM) features. At 28 days, DWI_original_glrlm_RunVariance, reflecting the spatial heterogeneity of the diffusion signal within the lesion,demonstrated the strongest association with mortality (HR: 2.43, $p < 0.005$), followed by CT_original_glszm_GrayLevelNonUniformity representing heterogeneity in CT attenuation within the lesion(HR: 1.75, $p < 0.005$), DWI_original_firstorder_Skewness describing the asymmetry of the diffusion signal intensity distribution: 1.50, $p = 0.015$), and CT_diagnostics_Image.original_Maximum indicating the highest CT attenuation within the lesion (HR: 1.50, $p = 0.017$). At 90 days, DWI_original_glrlm_RunVariance remained the strongest predictor of mortality (HR: 2.29, $p < 0.005$). Additional significant predictors included FLAIR_original_firstorder_TotalEnergy which indicates the overall signal energy within the lesion (HR: 1.71, $p < 0.005$), CT_original_glszm_GrayLevelNonUniformity (HR: 1.70, $p < 0.005$), CT_original_shape_MinorAxisLength representing the short-axis dimension of the lesion (HR: 1.72, $p = 0.005$), CT_diagnostics_Image.original_Maximum (HR: 1.53, $p = 0.008$), and DWI_original_firstorder_Skewness (HR: 1.39, $p = 0.049$).At 180 days, four radiomics features remained significantly associated with mortality, including DWI_original_glrlm_RunVariance (HR: 2.39, $p < 0.005$), CT_original_glszm_GrayLevelNonUniformity (HR: 1.74, $p < 0.005$), CT_original_shape_MinorAxisLength (HR: 1.80, $p < 0.005$), and CT_diagnostics_Image.original_Maximum (HR: 1.53, $p = 0.007$).At 365 days, DWI_original_glrlm_RunVariance continued to demonstrate the strongest association with mortality (HR: 2.33, $p < 0.005$), followed by CT_original_shape_MinorAxisLength (HR: 1.84, $p < 0.005$), CT_diagnostics_Image.original_Maximum (HR: 1.47, $p = 0.014$), DWI_original_firstorder_Skewness (HR: 1.43, $p = 0.020$), and FLAIR_original_firstorder_Kurtosis which characterizes the distribution of FLAIR signal intensities within the lesion(HR: 1.42, $p = 0.025$). Overall, texture-based radiomics features predominated among the significant predictors, with GLRLM and GLSZM features demonstrating the most consistent associations with mortality **(Supplementary Table 3)**.

**Supplementary Table 1.** Demographic and clinical characteristics of survivors and non-survivors at 28, 90, 180, and 365 days

| Variables | Survivors_28th Day | Non-Survivors_28th Day | p value | Survivors_90th Day | Non-Survivors_90th Day | p value |
|---|---|---|---|---|---|---|
| N | 37 | 26 | | 32 | 31 | |
| Age | 54.2 (13.1) | 63.2 (11.7) | 0.007[†] | 53.2 (13.6) | 62.9 (11.0) | 0.003[†] |
| Gender (Male) | 27 (73.0%) | 13 (50.0%) | 0.062[‡] | 24 (75.0%) | 16 (51.6%) | 0.054[‡] |
| Gender (Female) | 10 (27.0%) | 13 (50.0%) | | 8 (25.0%) | 15 (48.4%) | |
| Tobacco | 7 (18.9%) | 6 (23.1%) | 0.688[‡] | 7 (21.9%) | 6 (19.4%) | 0.805[‡] |
| Alcohol | 8 (21.6%) | 5 (19.2%) | 0.817[‡] | 8 (25.0%) | 5 (16.1%) | 0.384[‡] |
| Arterial Hypertension | 34 (91.9%) | 23 (88.5%) | 0.684[¶] | 29 (90.6%) | 28 (90.3%) | 1.000[¶] |
| Diabetes Mellitus | 15 (40.5%) | 13 (50.0%) | 0.457[‡] | 11 (34.4%) | 17 (54.8%) | 0.102[‡] |
| Dyslipidemia | 7(18.9%) | 6 (23.1%) | 0.688[‡] | 6 (18.8%) | 7 (22.6%) | 0.707[‡] |
| Previous Stroke | 9 (24.3%) | 11 (42.3%) | 0.131[‡] | 6 (18.8%) | 14 (45.2%) | 0.024[‡] |
| AF | 3 (8.1%) | 3 (11.5%) | 0.684[¶] | 2 (6.3%) | 4 (12.9%) | 0.426[¶] |
| CAD | 6 (16.2%) | 7 (26.9%) | 0.301[‡] | 5 (15.6%) | 8 (25.8%) | 0.318[‡] |
| CKD | 4 (10.8%) | 2 (7.7%) | 1.000[¶] | 4 (12.5%) | 2 (6.5%) | 0.672[¶] |

| Variables | Survivors_180th Day | Non-Survivors_180th Day | p value | Survivors_365th Day | Non-Survivors_365th Day | p value |
|---|---|---|---|---|---|---|
| N | 30 | 33 | | 25 | 37 | |
| Age | 53.0 (39.8-63.3) | 64.0 (59.0-69.5) | 0.002§ | 52.1 (13.9) | 61.5 (11.4) | 0.005† |
| Gender (Male) | 24 (80.0%) | 16 (48.5%) | 0.009‡ | 21 (84.0%) | 18 (48.6%) | 0.005‡ |
| Gender (Female) | 6 (20.0%) | 17 (51.5%) | | 4 (16.0%) | 19 (51.4%) | |
| Tobacco | 7 (23.3%) | 6 (18.2%) | 0.614‡ | 7 (28.0%) | 6 (16.2%) | 0.264‡ |
| Alcohol | 8 (26.7%) | 5 (15.2%) | 0.259‡ | 8 (32.0%) | 5 (13.5%) | 0.079‡ |
| Arterial Hypertension | 27 (90.0%) | 30 (90.9%) | 1.000¶ | 22 (88.0%) | 34 (91.9%) | 0.678¶ |
| Diabetes Mellitus | 11 (36.7%) | 17 (51.5%) | 0.236‡ | 10 (40.0%) | 18 (48.6%) | 0.502‡ |
| Dyslipidemia | 6 (20.0%) | 7 (21.2%) | 0.905‡ | 5 (20.0%) | 8 (21.6%) | 0.878‡ |
| Previous Stroke | 6 (20.0%) | 14 (42.4%) | 0.056‡ | 5 (20.0%) | 15 (40.5%) | 0.090‡ |
| AF | 2 (6.7%) | 4 (12.1%) | 0.674¶ | 0 (0.0%) | 6 (16.2%) | 0.073¶ |
| CAD | 5 (16.7%) | 8 (24.2%) | 0.458‡ | 2 (8.0%) | 11 (29.7%) | 0.039‡ |
| CKD | 4 (13.3%) | 2 (6.1%) | 0.412¶ | 3 (12.0%) | 2 (5.4%) | 0.385¶ |

*N, number of participants. Normally distributed continuous variables are presented as mean ± SD, and group comparisons were performed using an independent-samples t-test (†). Non-normally distributed continuous variables are presented as median (IQR), and group comparisons were performed using the Mann–Whitney U test (§). Categorical variables are presented as N (%). p values for categorical variables were calculated using Pearson's chi-square test (‡) or Fisher's exact test (¶). One participant was lost to follow-up at 365 days and was excluded from the 365-day outcome analysis.*

**Supplementary Table 2.** Clinical features obtained from univariate Cox regression analyses

| Features | HR (95% CI) | p value |
|---|---|---|
| **28 days** | | |
| NIHSS | 1.06 (1.03-1.10) | <0.005 |
| NLR | 1.04 (1.02-1.07) | <0.005 |
| Age (years) | 1.05 (1.01-1.08) | 0.011 |
| CRP (mg/L) | 1.01 (1.00-1.01) | 0.037 |
| **90 days** | | |
| NIHSS | 2.34 (1.61-3.41) | <0.005 |
| NLR | 1.53 (1.19-1.97) | <0.005 |
| CRP (mg/L) | 1.53 (1.14-2.06) | <0.005 |
| Age (years) | 1.81 (1.20-2.74) | <0.005 |
| Stroke Onset-to-Admission Time (days) | 1.57 (1.12-2.20) | 0.009 |
| Hemoglobin (gm/dl) | 0.64 (0.43-0.94) | 0.024 |
| LAMS | 1.61 (1.06-2.46) | 0.027 |
| Albumin (gm/dl) | 0.70 (0.50-0.98) | 0.037 |
| Gender | 0.48 (0.24-0.97) | 0.042 |
| **180 days** | | |
| NIHSS | 2.28 (1.58-3.27) | <0.005 |
| NLR | 1.52 (1.18-1.96) | <0.005 |
| Age (years) | 1.87 (1.25-2.80) | <0.005 |
| CRP (mg/L) | 1.56 (1.17-2.08) | <0.005 |
| Stroke Onset-to-Admission Time (days) | 1.61 (1.16-2.24) | <0.005 |
| Gender | 0.41 (0.21-0.82) | 0.012 |
| Hemoglobin (gm/dl) | 0.63 (0.43-0.92) | 0.017 |
| Albumin (gm/dl) | 0.70 (0.51-0.97) | 0.031 |
| LAMS | 1.54 (1.03-2.29) | 0.035 |
| Diagnosis | 2.17 (1.01-4.67) | 0.048 |
| **365 days** | | |
| NIHSS | 2.15 (1.53-3.01) | <0.005 |
| CRP (mg/L) | 1.58 (1.19-2.10) | <0.005 |
| Age (years) | 1.73 (1.20-2.50) | <0.005 |
| NLR | 1.46 (1.12-1.91) | 0.005 |
| Gender | 0.40 (0.21-0.76) | 0.005 |
| Albumin (gm/dl) | 0.64 (0.46-0.88) | 0.006 |
| Stroke Onset-to-Admission Time (days) | 1.57 (1.13-2.19) | 0.008 |
| Hemoglobin (gm/dl) | 0.61 (0.43-0.88) | 0.008 |
| Diagnosis | 2.54 (1.19-5.39) | 0.016 |

**Supplementary Table 3.** Radiomics features obtained from univariate Cox regression analyses

| Features | HR (95% CI) | p value |
|---|---|---|
| **28 days** | | |
| DWI_original_glrlm_RunVariance | 2.43 (1.69-3.48) | <0.005 |
| CT_original_glszm_GrayLevelNonUniformity | 1.75 (1.29-2.36) | <0.005 |
| DWI_original_firstorder_Skewness | 1.50 (1.08-2.09) | 0.015 |
| CT_diagnostics_Image.original_Maximum | 1.50 (1.08-2.09) | 0.017 |
| **90 days** | | |
| DWI_original_glrlm_RunVariance | 2.29 (1.61-3.24) | <0.005 |
| FLAIR_original_firstorder_TotalEnergy | 1.71 (1.27-2.30) | <0.005 |
| CT_original_glszm_GrayLevelNonUniformity | 1.70 (1.25-2.31) | <0.005 |
| CT_original_shape_MinorAxisLength | 1.72 (1.18-2.52) | 0.005 |
| CT_diagnostics_Image.original_Maximum | 1.53 (1.12-2.10) | 0.008 |
| DWI_original_firstorder_Skewness | 1.39 (1.01-1.93) | 0.049 |
| **180 days** | | |
| DWI_original_glrlm_RunVariance | 2.39 (1.69-3.38) | <0.005 |
| CT_original_glszm_GrayLevelNonUniformity | 1.74 (1.29-2.35) | <0.005 |
| CT_original_shape_MinorAxisLength | 1.80 (1.25-2.61) | <0.005 |
| CT_diagnostics_Image.original_Maximum | 1.53 (1.13-2.09) | 0.007 |
| **365 days** | | |
| DWI_original_glrlm_RunVariance | 2.33 (1.66-3.27) | <0.005 |
| CT_original_shape_MinorAxisLength | 1.84 (1.29-2.63) | <0.005 |
| CT_diagnostics_Image.original_Maximum | 1.47 (1.08-2.01) | 0.014 |
| DWI_original_firstorder_Skewness | 1.43 (1.06-1.94) | 0.020 |
| FLAIR_original_firstorder_Kurtosis | 1.42 (1.04-1.92) | 0.025 |
